# Incidence and prevalence of hidradenitis suppurativa across spondyloarthritis-related diseases

**DOI:** 10.1101/2025.03.02.25323167

**Authors:** Sizheng Steven Zhao, Uazman Alam, Qingfan Yang, Zhimin Liu, Zenas Yiu

**Author notes:** **Correspondence to:** Dr Sizheng S Zhao. Centre for Epidemiology Versus Arthritis, Division of Musculoskeletal and Dermatological Science, School of Biological Sciences, Faculty of Biological Medicine and Health, The University of Manchester, Manchester Academic Health Science Centre, Oxford Road, Manchester, M13 9LJ, UK.

## Abstract

**Importance:** Hidradenitis suppurativa (HS) has been associated with spondyloarthritis (SpA) and inflammatory bowel disease (IBD) and proposed as a potential manifestation of these conditions. However, the relative rarity of HS, combined with the diverse methodologies used in previous studies, leaves several uncertainties surrounding its epidemiology within this disease spectrum.

**Objective:** To describe the prevalence and incidence of HS across pathologically related diseases within the spondyloarthritis (SpA) family, namely psoriasis, psoriatic arthritis (PsA), axial spondyloarthritis (axSpA), uveitis, Crohn’s disease, and ulcerative colitis.

**Design:** Cohort study conducted using data between January 2005 and January 2025.

**Setting:** The study used data from electronic health records of over 135 million individuals across more than 100 healthcare organizations in North America.

**Participants:** Six disease populations ranging from 46,928 to 191,605 individuals.

**Exposures:** Populations of psoriasis, PsA, axial spondyloarthritis, uveitis, Crohn’s disease, and ulcerative colitis.

**Main outcomes and measures:** Prevalence and incidence per 100,000 patient-years of HS.

**Results:** The prevalence of HS was 0.4% in psoriasis and PsA, higher than axSpA (0.2%), Crohn’s disease (0.3%) or ulcerative colitis (0.2%). HS incidence was higher in psoriasis (124/100,000 patient-years) and PsA (119/100,000 patient-years) than other SpA-related disease (range 69 to 88/100,000 patient-years), with the exception of Crohn’s disease (177//100,000 patient-years).

**Conclusions and Relevance:** This study highlights a significant epidemiological association between HS and SpA-related diseases, particularly psoriatic disease and Crohn’s disease. Clinicians providing care to these patient groups should consider enquiring about HS symptoms, although overall incidence rates are low. Further research is needed into the shared pathophysiology of HS and psoriatic disease.

**Key points:** *Question:* What is the prevalence and incidence of hidradenitis suppurativa (HS) in conditions with shared immunopathology such as psoriatic disease, axial spondyloarthritis, uveitis, and inflammatory bowel disease?

*Findings:* This cohort study found high prevalence and incidence of HS among individuals with psoriatic disease and confirmed previously reported high incidence in Crohn’s disease. Incidence rates of HS in other diseases of the spondyloarthritis family, such as axial spondyloarthritis, were lower.

*Meaning:* Clinicians providing care for people with psoriatic disease should consider enquiring about HS symptoms, although overall incidence rates are low.

## Introduction

Hidradenitis suppurativa (HS) is a chronic inflammatory condition that has substantial impact on quality of life as well as both social and physical function. Estimates of HS prevalence vary widely, ranging from 0.05% to 4.1%, depending on methodology and geographic location [1]. HS has been associated with spondyloarthritis (SpA) and inflammatory bowel disease (IBD) and has even been proposed as a potential manifestation of these conditions [2]. In a cross-sectional survey of people with SpA in the Netherlands, HS prevalence was reported to be 9.1% [2]. In a population-based cohort study from the US, individuals with IBD were around 9 times more likely to develop HS than the general population [3]. These associations may be explained by shared pathoaetiology, for example, dysregulated TNF and/or IL-17 signalling, which aligns with therapeutic overlap observed across these conditions. Psoriatic disease is also associated with metabolic comorbidities that are strongly implicated in HS.

The relative rarity of HS compared to other SpA-related diseases presents challenges for research, leading to several uncertainties regarding its epidemiology. First, whether HS is specifically associated with SpA-related diseases remains unclear, as it has also been linked to rheumatoid arthritis (RA) [4], which has different pathoaetiology [5]. Second, it is uncertain whether HS is associated with all members of the SpA-related disease or only a subset; for example, HS appears more common in Crohn’s disease than ulcerative colitis [6], but it is unclear whether its association with SpA reflects axial SpA or psoriatic arthritis (PsA). Third, comparing the relative incidence of HS across SpA-related diseases is complicated by differing methodologies across studies [1].

Better understanding HS epidemiology in SpA-related diseases is essential for mechanistic understanding, screening and early diagnosis, planning healthcare provision, and identifying potential opportunities for drug repurposing. Our study aimed to compare the incidence and prevalence of HS across pathologically related diseases within the SpA family, specifically psoriasis, psoriatic arthritis (PsA), axial spondyloarthritis, uveitis, Crohn’s disease, ulcerative colitis, and, as comparators, seropositive and seronegative RA.

## Methods

### Data source

We conducted a cohort study using data from electronic health records of over 135 million individuals across more than 100 healthcare organizations (HCOs), mostly secondary and tertiary centres in North America. Further details regarding the database are available at trinetx.com and have been previously described [7]. Analyses are performed at individual HCOs with only aggregate results returned to the platform. Additional ethical approval was not required because this study used only de-identified data and did not involve the collection, use, or transmittal of identifiable data.

### Study Population and outcome

We defined the following populations using individuals aged 18 or older with at least two ICD codes for the index disease: psoriatic arthritis (L40.5) excluding any codes for ankylosing spondylitis (M45), rheumatoid arthritis (M05, M06) or enteropathic arthritis (M07); cutaneous only psoriasis vulgaris, henceforth referred to as psoriasis (L40.0 excluding L40.5); axial spondyloarthritis (M45 excluding L40.5, M05-M07); uveitis (H20); Crohn’s disease (K50, excluding K51); ulcerative colitis (K51, excluding K50). We included seronegative RA (M06, excluding M05, L40.5, M45) and seropositive RA (M05, excluding M06, L40.5, M45) as control outcomes. HS was defined using any occurrence of code L73.2.

For each disease population we describe the following factors relevant to HS risk: age at first ICD code for the index disease, gender, ethnicity (white versus non-white), type 2 diabetes (E11), hypertension (I10), chronic obstructive pulmonary disease (COPD) and emphysema (J44, J43) as proxies for smoking, polycystic ovarian syndrome (E28.2), BMI (kg/m^2^) and CRP (mg/L).

### Statistical Analysis

We reported the prevalence of HS at or before the date of the first ICD code for the index disease. The 95% confidence interval (CI) for prevalence was derived using P ± 1.96*√(P*(1-P)/n), where P is the prevalence. Incidence rate was calculated as events per 100,000 person-years, excluding individuals with prior history of HS. The 95% CI was approximated using Byar’s Poisson distribution for rare events E/PY*exp(±1.96/√E), where E is the number of events and PY the person-years. Follow-up started a month after the first ICD code for the index disease to reduce reverse causation and continued until the date of the last health record entry. Analyses were conducted using R, version 4.2.2, integrated into the TriNetX platform.

## Results

We identified eight disease populations ranging from 38,568 to 203,335 individuals (Table 1). The mean age was generally younger in SpA-related diseases than RA. The ratio of males-to-females was generally comparable across SpA-related diseases, except axSpA (61% male) and RA (24%).

**Table 1.** Baseline characteristics of spondyloarthritis-related diseases and rheumatoid arthritis as comparators.

|  | Psoriatic arthritis | Psoriasis | Axial SpA | Uveitis | Crohn's disease | Ulcerative colitis | Seronegative RA | Seropositive RA |
| --- | --- | --- | --- | --- | --- | --- | --- | --- |
| n | 77,504 | 114,347 | 46,928 | 111,513 | 191,605 | 174,839 | 384,208 | 38,261 |
| Age | 51.8 (15.6) | 48.7 (19.1) | 47.2 (17.9) | 48.3 (21.0) | 42.4 (19.8) | 48.7 (20.0) | 60.0 (16.3) | 59.2 (15.1) |
| Male | 31966 (41.2) | 51771 (45.3) | 28442 (60.6) | 48242 (43.3) | 82539 (43.1) | 77966 (44.6) | 93838 (24.4) | 9107 (23.8) |
| White | 54947 (70.9) | 63169 (55.2) | 22528 (48.0) | 51282 (46.0) | 127659 (66.6) | 113514 (64.9) | 234468 (61) | 18648 (48.7) |
| Overweight/obesity | 9259 (11.9) | 12224 (10.7) | 3152 (6.7) | 11331 (10.2) | 8533 (4.5) | 11382 (6.5) | 38985 (10.1) | 3221 (8.4) |
| Type 2 diabetes | 7499 (9.7) | 11258 (9.8) | 3082 (6.6) | 14318 (12.8) | 7661 (4.0) | 12290 (7.0) | 42435 (11) | 3122 (8.2) |
| Hypertension | 16334 (21.1) | 24718 (21.6) | 7059 (15) | 27216 (24.4) | 19217 (10) | 29786 (17) | 99199 (25.8) | 7640 (20) |
| COPD | 2139 (2.8) | 4034 (3.5) | 1154 (2.5) | 3566 (3.2) | 3514 (1.8) | 5329 (3) | 19722 (5.1) | 1406 (3.7) |
| Emphysema | 605 (0.8) | 1184 (1) | 313 (0.7) | 1212 (1.1) | 970 (0.5) | 1873 (1.1) | 5516 (1.4) | 484 (1.3) |
| PCOS | 533 (0.7) | 476 (0.4) | 186 (0.4) | 364 (0.3) | 548 (0.3) | 487 (0.3) | 1426 (0.4) | 90 (0.2) |
| BMI (kg/m<sup>2</sup>) | 31.5 (7.8) | 30.0 (7.8) | 29.1 (7.2) | 28.7 (7.4) | 27.2 (7.3) | 27.7 (6.7) | 29.9 (7.7) | 29.8 (7.4) |
| C reactive protein (mg/l) | 14.3 (31.7) | 19.0 (41.1) | 15.7 (32.6) | 18.6 (39.9) | 26.3 (44.4) | 24.6 (46.7) | 21.0 (41.5) | 18.6 (36.2) |
| Hidradenitis suppurativa | 281 (0.4) | 515 (0.5) | 99 (0.2) | 314 (0.3) | 538 (0.3) | 289 (0.2) | 857 (0.2) | 71 (0.2) |

BMI, body mass index; PCOS, polycystic ovary syndrome; COPD, chronic obstructive pulmonary disease.

The prevalence of HS at baseline (i.e., the date of the first ICD code for the index disease) was lowest for RA and ulcerative colitis (0.16-0.21%) and highest for PsA (0.35%; 95%CI 0.31-0.39%) and psoriasis (0.38%; 95%CI 0.35-0.42%) (Figure 1).

**Figure 1.**
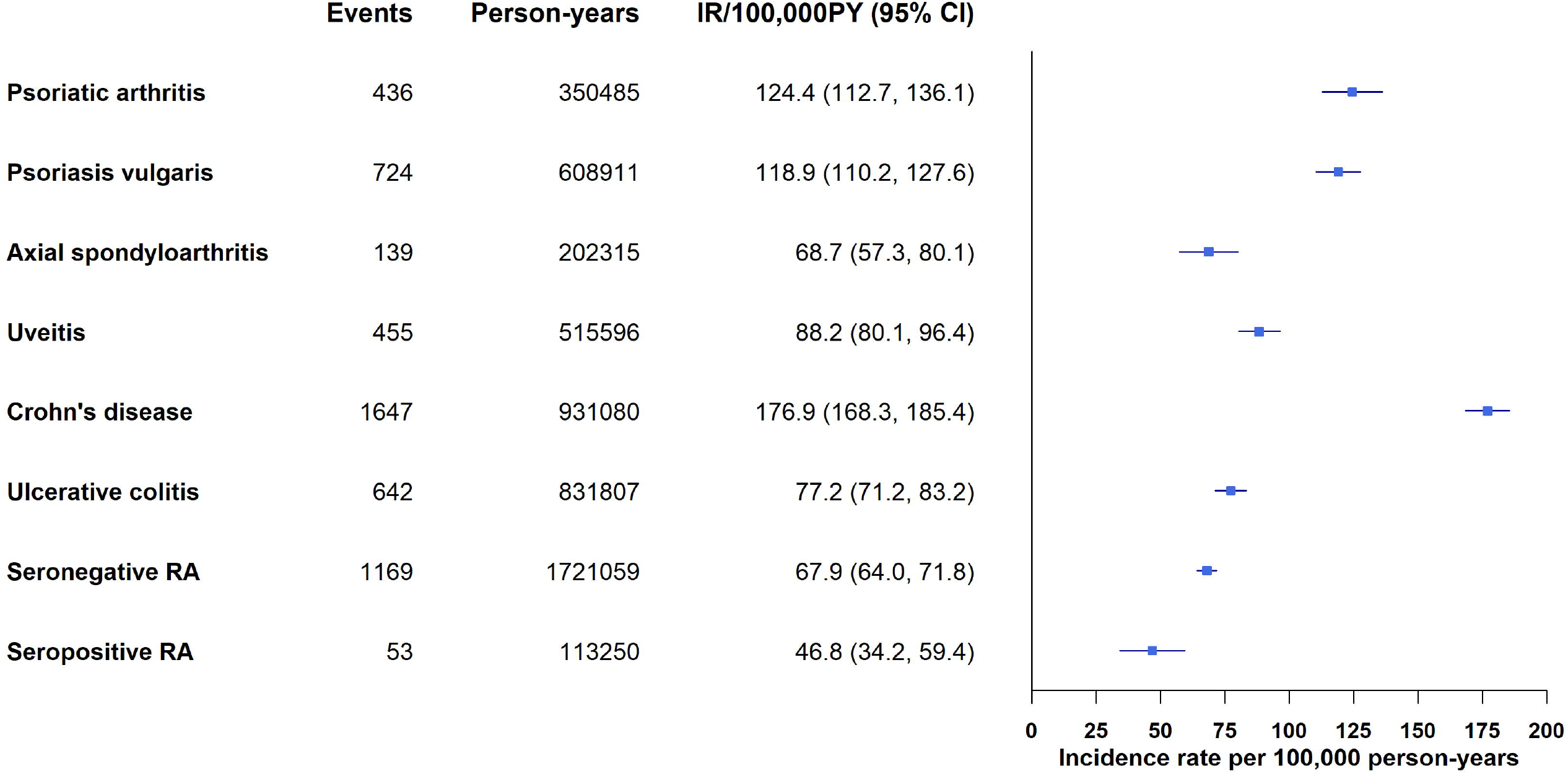
Prevalence of hidradenitis suppurativa at time of spondyloarthritis-related disease diagnosis. Legend: Events, number of HS cases; N, total sample size; CI confidence interval; RA, rheumatoid arthritis.

Incidence rate per 100,000 person-years was highest for Crohn’s disease (177; 95%CI 168, 185), then PsA (124; 95%CI 113, 136) and psoriasis (119; 95%CI 110, 128). Incidence rates were lower for axial spondyloarthritis, uveitis, UC (range 69-88 per 100,000 person-years), and lowest for seropositive RA (Figure 2).

**Figure 2.**
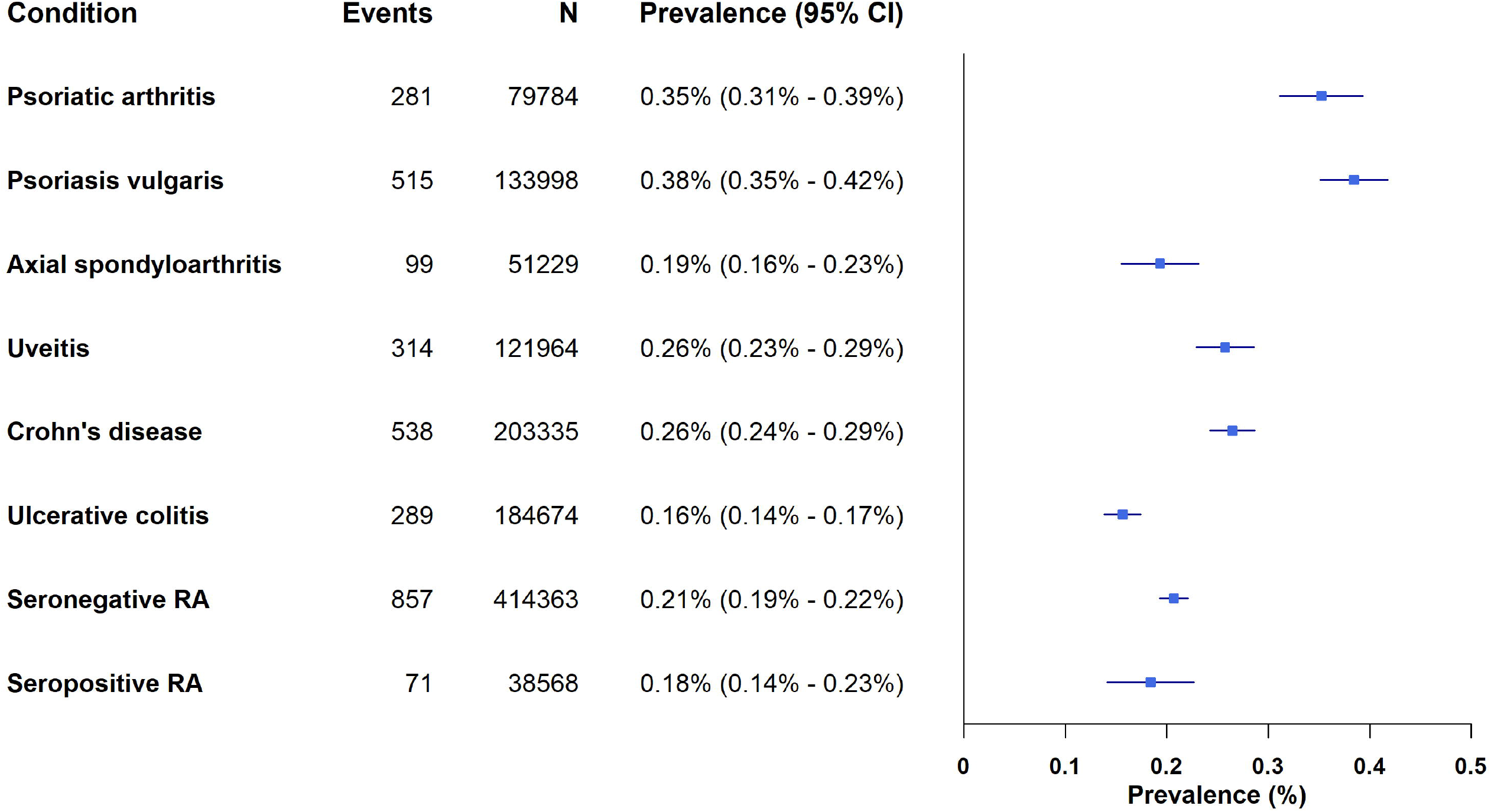
Incidence of hidradenitis suppurativa across spondyloarthritis-related diseases. Legend: Events, number of HS cases; N, total sample size; IR, incidence rate; CI confidence interval; RA, rheumatoid arthritis.

Events, number of HS cases; N, total sample size; CI confidence interval; RA, rheumatoid arthritis. Date of the first ICD code for the index disease was used to approximate its diagnosis.

## Discussion

In this study of HS epidemiology across spondyloarthritis-related diseases, we observed a notably high incidence and prevalence of HS among individuals with psoriatic disease. The prevalence of HS was higher in psoriatic disease compared to IBD, axial spondyloarthritis, or uveitis at the time of diagnosis. Similarly, HS incidence was higher in psoriatic disease than in axial spondyloarthritis, uveitis, or ulcerative colitis, with the highest incidence observed in Crohn’s disease. These findings support an epidemiological association between HS and SpA-related diseases, particularly psoriatic and Crohn’s disease.

Most prior research on HS epidemiology has focused on population estimates, which vary widely, with estimates from Western countries differing by nearly 100-fold (0.05% to 4.1%) [1]. In the US, population estimates ranged from 0.05-0.1% [8,9], which provides context for our results where prevalence estimates were generally higher (except in seropositive RA). The survey-estimate of 9.1% prevalence in a Dutch axSpA population [2] is likely an overestimate due to use of screening questions and incomplete survey response. Our findings suggest that both prevalence and incidence of HS are lower in axial spondyloarthritis than in PsA, uveitis or Crohn’s. Studies of HS incidence are scarce, with population estimates of 6 to 11.4/100,000PY in the US [10,11], which although not directly comparable again supports higher incidence in SpA related diseases reported herein.

Our results confirm previous associations between HS and Crohn’s disease but indicate that the link between HS and axial spondyloarthritis is less pronounced than with psoriatic disease. The stronger association with psoriatic disease may stem from a combination of shared inflammatory dysfunction and risk factors, particularly metabolic syndrome. These findings support the epidemiological association with the SpA family of diseases, although further research into the genetics of HS is needed to clarify these relationships.

A strength of this study is our use of standardised methodologies across SpA-related diseases within a large, non-selective data source. However, several limitations exist. First, we could not compare our findings to a population representative of the general population, as our data source was based on electronic health records. Second, ICD-based disease definitions have imperfect accuracy, with underdiagnosis being a significant concern. Nonetheless, our estimates were generally higher than prior US population estimates, which is reassuring. Lastly, differences in HS prevalence and incidence may partly reflect variations in demographics and clinical characteristics across SpA-related diseases. However, addressing these differences was not the focus of the current study.

In conclusion, this study highlights high incidence and prevalence of HS in psoriatic disease and confirms its high incidence in Crohn’s disease. Clinicians providing care to these patient groups should consider enquiring about HS symptoms, although overall incidence rates are low. Further research is needed into the shared pathophysiology of HS and psoriatic disease.

## Data Availability

The data that support the findings of this study are available from TriNetX, LLC but third-party restrictions apply to the availability of these data. The data were used under license for this study with restrictions that do not allow for the data to be redistributed or made publicly available. However, for accredited researchers, the TriNetX data is available for licensing at TriNetX, LLC. To gain access to the data in the TriNetX research network, a request can be made to TriNetX (trinetx.com), but costs may be incurred, a data sharing agreement would be necessary, and no patient identifiable information can be obtained.

https://trinetx.com/

## Acknowledgements

We thank TriNetX (TriNetX LLC, Cambridge, MA, USA) for access to their federated network.

## Author contributions

Dr Zhao had full access to all the data in the study and takes responsibility for the integrity of the data and the accuracy of the data analysis.

## Data Sharing Statement

The data that support the findings of this study are available from TriNetX, LLC but third-party restrictions apply to the availability of these data. The data were used under license for this study with restrictions that do not allow for the data to be redistributed or made publicly available. However, for accredited researchers, the TriNetX data is available for licensing at TriNetX, LLC. To gain access to the data in the TriNetX research network, a request can be made to TriNetX (trinetx.com), but costs may be incurred, a data sharing agreement would be necessary, and no patient identifiable information can be obtained. No data from Liverpool University Hospitals NHS Foundation Trust was utilized in this analysis.

## References

1. Bukvić Mokos Z, Markota Čagalj A, Marinović B. Epidemiology of hidradenitis suppurativa. Clin. Dermatol. 2023;41:564–75.

2. Rondags A, Arends S, Wink FR, Horváth B, Spoorenberg A. High prevalence of hidradenitis suppurativa symptoms in axial spondyloarthritis patients: A possible new extra-articular manifestation. Semin. Arthritis Rheum. 2019;48:611–7.

3. Yadav S, Singh S, Edakkanambeth Varayil J, Harmsen WS, Zinsmeister AR, Tremaine WJ, et al. Hidradenitis Suppurativa in Patients With Inflammatory Bowel Disease: A Population-Based Cohort Study in Olmsted County, Minnesota. Clin. Gastroenterol. Hepatol. Off. Clin. Pract. J. Am. Gastroenterol. Assoc. 2016;14:65–70.

4. Kridin K, Shavit E, Damiani G, Cohen AD. Hidradenitis suppurativa and rheumatoid arthritis: evaluating the bidirectional association. Immunol. Res. 2021;69:533–40.

5. McGonagle D, Aydin SZ, Gül A, Mahr A, Direskeneli H. ‘MHC-I-opathy’—unified concept for spondyloarthritis and Behçet disease. Nat. Rev. Rheumatol. 2015;11:731–40.

6. Chen WT, Chi CC. Association of Hidradenitis Suppurativa With Inflammatory Bowel Disease: A Systematic Review and Meta-analysis. JAMA Dermatol. 2019;155:1022–7.

7. Zhao SS, Riley D, Hernandez G, Alam U. Comparative safety of JAK inhibitors versus TNF or IL-17 inhibitors for cardiovascular disease and cancer in psoriatic arthritis and axial spondyloarthritis. Clin. Ther. [Internet] 2025 [cited 2025 Jan 14];Available from: https://research.manchester.ac.uk/en/publications/comparative-safety-of-jak-inhibitors-versus-tnf-or-il-17-inhibito

8. Cosmatos I, Matcho A, Weinstein R, Montgomery MO, Stang P. Analysis of patient claims data to determine the prevalence of hidradenitis suppurativa in the United States. J. Am. Acad. Dermatol. 2013;68:412–9.

9. Garg A, Kirby JS, Lavian J, Lin G, Strunk A. Sex- and Age-Adjusted Population Analysis of Prevalence Estimates for Hidradenitis Suppurativa in the United States. JAMA Dermatol. 2017;153:760–4.

10. Garg A, Lavian J, Lin G, Strunk A, Alloo A. Incidence of hidradenitis suppurativa in the United States: A sex- and age-adjusted population analysis. J. Am. Acad. Dermatol. 2017;77:118–22.

11. Vazquez BG, Alikhan A, Weaver AL, Wetter DA, Davis MD. Incidence of hidradenitis suppurativa and associated factors: a population-based study of Olmsted County, Minnesota. J. Invest. Dermatol. 2013;133:97–103.

